# BMI and future risk for COVID-19 infection and death across sex, age and ethnicity: preliminary findings from UK biobank

**DOI:** 10.1101/2020.06.05.20122226

**Authors:** Naveed Sattar, Frederick K Ho, Jason MR Gill, Nazim Ghouri, Stuart R Gray, Carlos A Celis-Morales, S Vittal Katikireddi, Colin Berry, Jill P Pell, John JV McMurray, Paul Welsh

## Abstract

We examined the link between BMI and risk of a positive test for SARS-CoV-2 and risk of COVID-19-related death among UK Biobank participants. Among 4855 participants tested for SARS-CoV-2 in hospital, 839 were positive and of these 189 died from COVID-19. Poisson models with penalised thin plate splines were run relating exposures of interest to test positivity and case-fatality, adjusting for confounding factors. BMI was associated strongly with positive test, and risk of death related to COVID-19. The gradient of risk in relation to BMI was steeper in those under 70, compared with those aged 70 years or older for COVID-19 related death (Pinteraction=0.03). BMI was more strongly related to test positivity (P_interaction_=0.010) and death (P_interaction_=0.002) in non-whites, compared with whites. These data add support for adiposity being more strongly linked to COVID-19-related deaths in younger people and non-white ethnicities. If future studies confirm causality, lifestyle interventions to improve adiposity status may be important to reduce the risk of COVID-19 in all, but perhaps particularly, non-white communities.

---

We recently proposed multiple mechanisms by which obesity/ectopic fat might enhance risk for severe COVID-19.^1^ Our work was stimulated by observations in hospital intensive care units which showed that body mass indices (BMI) above 35 or 40 were associated with poorer prognosis.^2,3^ However, whether BMI is linearly associated with COVID-19-related risk in both sexes, in non-white ethnicities, and across ages, is not known. Whether adiposity might mediate higher COVID-19-related risks, including death, in Black and other ethnic minority groups in the UK is important to understand. To address these issues, we used UK Biobank data to examine the association between BMI and test positivity for SARS-CoV-2 infection in hospital, as well as COVID-19-related deaths.

Over 500,000 participants (aged 50-81 on 16^th^ March 2020) from the UK general population were recruited between 2006-2010, with details described elsewhere. For this study, only participants from English assessment centres who were alive on 1^st^ March 2020 were included. Participants with complete data for baseline anthropometric variables and covariates were included (n=374,922). The exposure of interest was BMI. Public Health England (PHE) provided data on SARS-CoV-2 tests, including the specimen date, location, and result. Data were available for the period 16^th^ March 2020 to 31^st^ May 2020. Tests were conducted on 5,274 participants in the available cohort. Of these, 419 were excluded because the tests were conducted outside the hospital setting. Overall, 839 participants tested positive in hospital and 189 of these individuals died from COVID-19 out of 374,503 participants. Deaths were defined as COVID-19 related if there was an ICD-10 code U07.1 or U07.2 on the death certificate.

Robust Poisson models with penalised thin plate splines were run relating exposures of interest to positive tests in hospital. Models were adjusted for age, socioeconomic status (Townsend Index), ethnicity, smoking (current, former, never), alcohol intake (unit/week), and baseline cardiovascular disease and diabetes. Interactions between adiposity measures and three moderators (sex, current age [<70 vs ≥70], and ethnicity [white vs non-white], were tested.

BMI was strongly associated with SARS-CoV-2 test positivity, in a broadly linear fashion (**Figure 1a**), despite the model allowing for non-linear associations. There was a hint of a stronger gradient of risk across BMI (P_interaction_=0.09) in men compared with women for a positive SAR-SCoV-2 test, although no clear evidence for such interaction for COVID-19-related death (**Figure 1b**). There was a stronger gradient (P_interaction_=0.03) of risk across BMI for COVID-19-related deaths but not SARS-CoV-2 test positivity in those under 70 versus 70 years or above (**Figure 1c, d**). BMI was more strongly related to risk for SARS-CoV-2 test positivity (P_interaction_=0.010) and COVID-19-related death (P_interaction_=0.002) in non-whites versus whites (**Figures 1e, f**).

**Figure 1.**
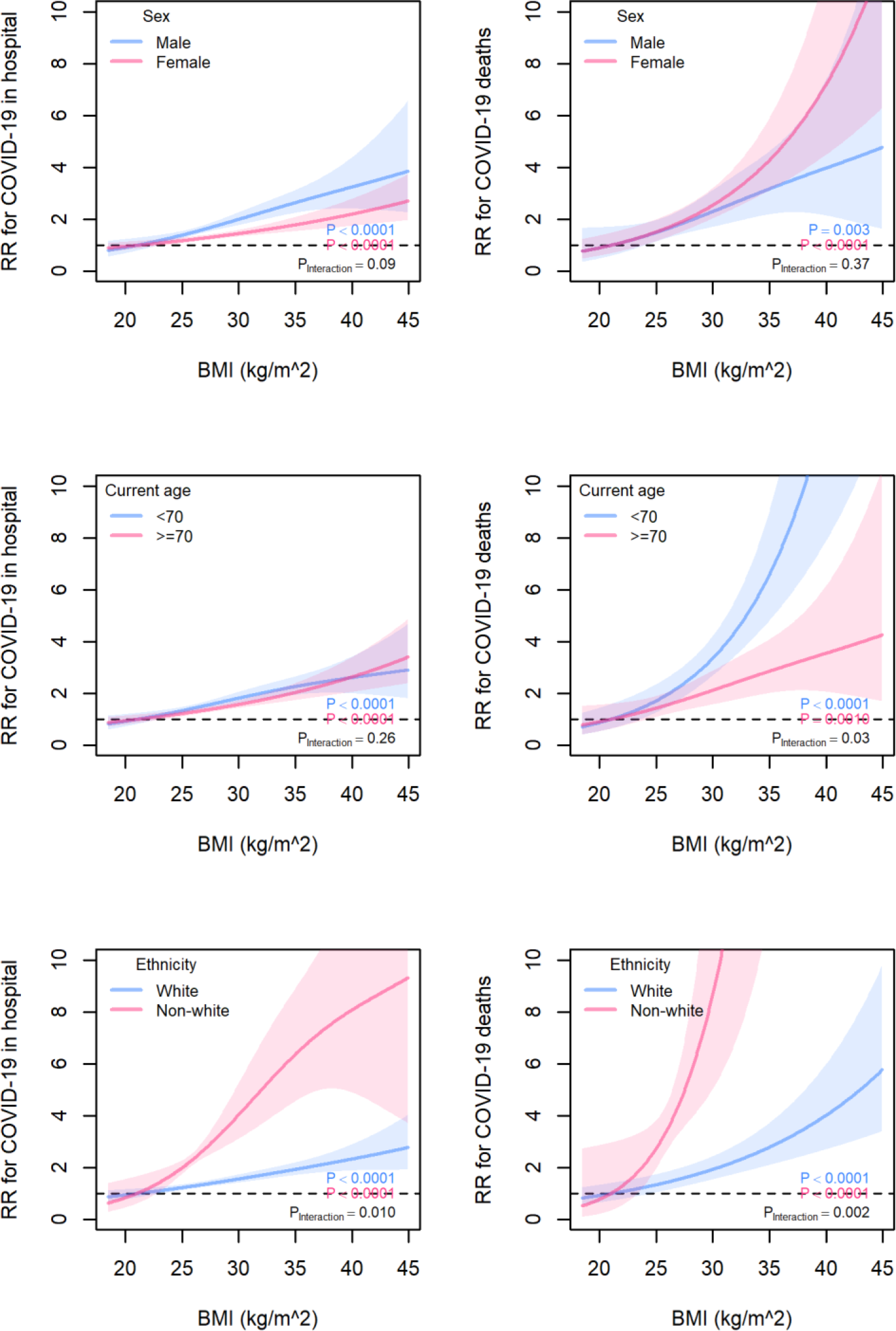
Associations of BMI with COVID-19 confirmed in hospital and COVID-19 deaths by sex (panels a to b), by age (panel c-d) and by ethnicity (panel e-f).

Our findings have some limitations. Only a small proportion of overall UK Biobank participants were tested for SARS-CoV-2 but consistent findings for test positivity and COVID-19-related death provide confidence that observations are likely real. Total numbers of deaths were also modest at 189. Those untested, as well as those tested negative, were grouped together in the analysis on the assumption that those tested in hospital were more likely to have severe disease. Baseline anthropometric measures were collected a median of 10.9 (IQR 9.7-12.4) years before SARS-CoV-2 testing was conducted. BMI levels, however, track well over time; the rank correlation for BMI was r=0.90 in 34, 917 repeat measurements taken 8.5 years post baseline, suggesting baseline BMI values reliably estimate BMI variance. We had modest numbers of deaths in subgroups in our analysis, requiring us to categorise race crudely as white and non-white. Associations with deaths are therefore less reliable. Whilst we adjusted for deprivation, individuals with higher weights may have had greater exposure to the virus. However, as excess fat can impair lung and metabolic function, stresses inflammatory pathways and cardiorenal systems, and promotes thrombotic responses,^1^ our findings likely reflect some causal elements. Although not formally tested, the current data suggest that BMI may be more strongly related to COVID-19 deaths than SARS-CoV-2 test positivity (**Figures 1a-f**).

Currently the UK government advises people with BMI>40 to be more stringent at adhering to non-pharmaceutical interventions (i.e. physical distancing, hand hygiene etc). Our results suggest that there is no evidence of a threshold effect at BMI=40, and that risk associations are linear. Moreover, our findings suggest excess fat may be a stronger risk factor for susceptibility to infection with SARS-CoV-2 and severity of resultant COVID-19, including death related to COVID-19, in non-whites compared to whites. Indeed, the gradients relating BMI to SARS-CoV-2 test positivity/COVID-19-related death in non-whites are reminiscent of the much stronger associations between BMI and diabetes associations in non-white people.^4^ This suggests, but does not fully prove, metabolic factors may be pertinent to excess COVID-19 severity in non-white populations, an observation in line with our suggested role for excess (ectopic) fat in COVID-19 severity.^1^ The stronger association of BMI with COVID-19-related death in younger individuals also makes sense given BMI is less precise measure of excess fat in elderly people due to changes in muscle mass linked to unintentional weight loss.

When taken in context with other emerging data,^2,3,5^ our results support the need for public health campaigns to help and support individuals across different ethnic groups who are overweight or obese to lose weight to lessen their chances of severe COVID-19. Such advice may be particularly pertinent to non-white communities. Whilst we lack trial evidence to prove weight loss lowers risks, there are many additional benefits of weight loss, including improved cardiorespiratory fitness and quality of life. We recognise that not all individuals are able to lose weight through lifestyle changes; some may require drug therapies, if even temporarily, to help reduce weight and maybe lower risk.

## Data Availability

All data that generated our work is available from the UK biobank and can be replicated.

## Sources of Funding

NS, JMRG, SRG, CACM, JJVM and PW acknowledge relevant research support from a British Heart Foundation Research Excellence Award (RE/18/6/34217), and from MMI - Kuwait and Dasman Diabetes Institute, Kuwait. SVK acknowledges funding from an NRS Senior Clinical Fellowship (SCAF/15/02), the Medical Research Council (MC_UU_12017/13) and the Scottish Government Chief Scientist Office (SPHSU13).

## Disclosures

NS reports personal fees from Amgen, AstraZeneca, Eli Lilly, Novo Nordisk, Pfizer, and Sanofi, and grants and personal fees from Boehringer Ingelheim, outside the submitted work. PW reports grant from Boehringer Ingelheim outside submitted work. All other authors declare no competing interests.

